# Pleural fluid analysis in neutropenia

**DOI:** 10.1101/2021.08.11.21261897

**Authors:** Isadore M. Budnick, Samuel F. Oliver, Andrew J. Barros, Jeffrey M. Sturek

## Abstract

Light’s criteria is the standard for determining the etiology of pleural effusions. Test characteristics of Light’s criteria varies in different populations, and Light’s criteria may misclassify effusions in patients with impaired immunity such as in neutropenia. This study evaluates the test characteristics of Light’s criteria and other clinically available pleural fluid tests in a cohort of patients with neutropenia. Cases were defined as a thoracentesis performed with an absolute neutrophil count of less than 1000 cells/µl documented at least 24 hours prior to the procedure. The etiology of the effusion was adjudicated by independent review of the case data and hospital course by two board certified pulmonologists. Categories for final diagnosis included exudate due to infection, exudate due to malignancy, exudate due to other, and transudate. A total of 83 thoracenteses from 80 patients were identified. Comorbidities included hematologic and solid tumor malignancies, recipients of allogeneic stem cell and solid organ transplants, heart failure, chronic kidney disease, and decompensated cirrhosis. Light’s criteria had a sensitivity of 92% and a specificity of 55% for identifying exudates, LR+ 2.07 and LR-0.14. A pleural fluid protein value of >2.9 g/dl had a sensitivity of 42% and a specificity of 96%, LR+11.12 and LR-0.61. When comparing exudative effusions, the percentage of neutrophils in the pleural fluid was significantly higher in infection, despite peripheral neutropenia. Together these results show a reduced specificity of Light’s criteria in neutropenia, and underscore the complexity of pleural effusions in this setting.

---

Light’s criteria remains the standard for pleural fluid analysis^1^. Light’s criteria has superb sensitivity but less robust specificity, particularly in certain populations^2^. Hematopoietic stem cell transplant recipients and patients with hematologic malignancies often develop pleural effusions^3,4^. Cohort studies within these populations have used Light’s criteria as the reference standard for case adjudication; however, the majority of effusions were classified as transudate or exudate without a specific etiology^3,4,5,6^. Light’s criteria may misclassify effusions in patients with neutropenia and lead to difficulties with identifying causative etiologies. This study examines the test characteristics of Light’s criteria and other pleural fluid tests that help to identify exudative effusions in a cohort of patients with neutropenia.

## Methods

Study was approved by UVA Health-Sciences Research IRB with approval number 22067. This study is retrospective with a waiver of patient consent. This cohort was created by querying the University of Virginia data warehouse for thoracenteses performed between March 2011 and June 2020 on patients who were neutropenic at least 24 hours prior to the thoracentesis. Neutropenia was defined by an absolute neutrophil count of less than 1000 cells/µl. All etiologies of neutropenia and indications for thoracentesis were included in this study. Clinical data, pleural fluid data, serum lab data, and imaging studies were extracted from the electronic medical record and stored in a REDCap database^7,8^.

### Case Adjudication

A summary of the index hospitalization was generated by authors IB and SO. Two board certified pulmonologists (AB and JS) independently reviewed the available data and case summary to determine the etiology of the effusion. After the independent review, the independent outcomes were compared and a consensus outcome was obtained. Categories for final diagnosis included exudate due to infection, exudate due to malignancy, exudate due to other, and transudate. Case definitions were established prior to case adjudication and were based on review of available clinical data as outlined in prior studies^9^.

### Data analysis

Determining if an effusion was an exudate was the outcome of interest. Case adjudication was the reference standard. Sensitivity, specificity, positive likelihood ratio (LR+), and negative likelihood ratio (LR-) were determined for each of the following: Light’s criteria, two-test rule, three-test rule, and the individual components of these tests as previously described^1,9^. Analysis of pleural fluid cholesterol was not completed due to the limited number of samples in this cohort.

Pleural fluid data from the exudates due to infection and exudates due to malignancy were compared. The variables of interest included pleural fluid values of pH, LDH, protein, glucose, total cell count, and cell count differential. Mean values and standard deviations were calculated for these variables. An independent sample t-test assuming unequal variances was used for comparison of mean values. Two-tail p values were reported with each variable of interest. A Bonferroni correction was performed to correct for multiplicity with a familywise error rate of 0.05.

## Results

### Baseline demographics

The search query identified 83 thoracenteses from 80 patients. The median age was 61.6 years old and 48.8% were male. The cohort was comprised of individuals with hematologic and solid tumor malignancies, including acute or chronic leukemia (26.3%), lymphoma of unspecified subtype (18.8%), non-pulmonary solid tumors (21.3%), and pulmonary solid tumors (13.8%). The cohort also included recipients of allogeneic stem cell transplants (11.3%), recipients of solid organ transplants (11.3%), individuals with heart failure (12.0%), chronic kidney disease (7.2%), and decompensated cirrhosis (8.4%). Duration of neutropenia prior to thoracentesis varied within the cohort. 30.1% of thoracenteses occurred with neutropenia of less than 48 hours, 15.7% with neutropenia of less than 7 but greater than 2 days, 28.9% with neutropenia of less than 30 but greater than 7 days, and 25.3% with neutropenia of greater than 30 days. The pleural effusions were determined by case adjudication to be transudates in 33.7% cases, exudates due to infection in 37.3%, exudates due to malignancy in 25.3%, and exudates due to other etiology in 3.6%.

### Test characteristics

Light’s criteria had a sensitivity of 92% and specificity of 55% for determining if the effusion was an exudate with an associated LR+ 2.07 and LR-0.14 (Table). Pleural fluid/serum protein ratio > 0.5 had a sensitivity of 58% and specificity of 89% with an associated LR+ 5.22 and LR-0.47 (Table). Pleural fluid protein values > 2.9 g/dl had a sensitivity of 42% and a specificity of 96% with a corresponding LR+ 11.12 and LR-0.61 (Table). All other tests had inferior test characteristics (Table).

**Table.** Neutropenic Pleural Fluid Test Characteristics

| Parameter | Sample size, n | Sensitivity, % | Specificity, % | LR+ | LR- |
| --- | --- | --- | --- | --- | --- |
| Light's criteria | 78 | 92 | 55 | 2.07 | 0.14 |
| Pleural fluid protein/serum ratio > 0.5 | 77 | 58 | 89 | 5.22 | 0.47 |
| Pleural fluid LDH/serum LDH ratio > 0.6 | 67 | 68 | 59 | 1.67 | 0.54 |
| Pleural fluid LDH > $\frac{2}{3}$ ULN | 76 | 60 | 73 | 2.23 | 0.55 |
| Two-test rule | 77 | 84 | 38 | 1.37 | 0.41 |
| Three-test rule | 77 | 90 | 40 | 1.51 | 0.24 |
| PF Protein > 2.9 | 78 | 42 | 96 | 11.12 | 0.61 |
| PF LDH > 0.45 ULN | 76 | 83 | 42 | 1.42 | 0.42 |

### Exudative effusions

Pleural fluid studies from the exudates due to infection versus exudates due to malignancy were compared to determine significant differences. Percentage of neutrophils within the pleural fluid cell count was significantly higher in the exudate due to infection as compared to exudate due to malignancy (Figure). None of the other variables of interest had a statistically significant difference between groups (Figure).

**Figure.**
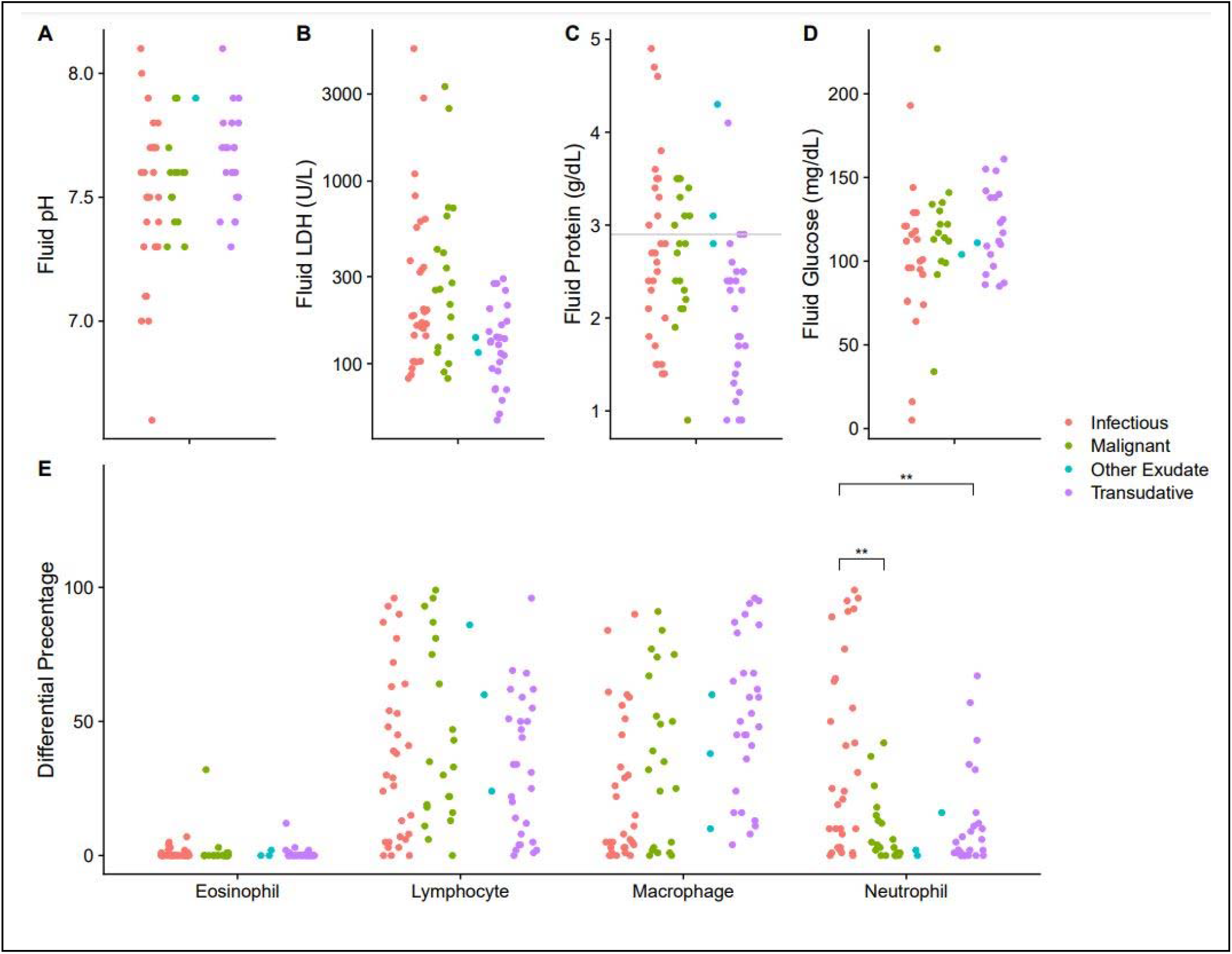
Neutropenic Pleural Fluid Laboratory Results. A-D, Comparison of fluid pH, LDH, protein, and glucose, respectively, between all four case categories. Horizontal line in C represents pleural fluid protein 2.9 as referenced in Table. E, Comparison of fluid cell profile between all four case categories. ** *P* value determined by Bonferroni correction with familywise error rate of 0.05.

## Discussion

This study is the first to analyze the test characteristics of Light’s criteria in a cohort of neutropenic patients using the clinical diagnosis as the reference standard. Light’s criteria demonstrated similar sensitivity but worse specificity compared to prior studies^2,10^. Light’s criteria specificity could suffer due to nonspecific protein and LDH elevations in inflammatory chronic conditions wherein neutropenia is common^10^. Pleural fluid protein > 2.9 g/dl and pleural fluid/serum protein > 0.5 had robust LR+ in this population, which is similar to other studies^7^. Pleural fluid neutrophil percentage distinguished exudate due to infection versus malignancy despite the presence of neutropenia. Strengths of this study include independent case adjudication and clinical diagnosis as case definition. Limitations include cohort size and variable etiologies and duration of neutropenia. Future studies could utilize a similar approach but focus on a particular etiology of neutropenia.

## Data Availability

Clinical data, pleural fluid data, serum lab data, and imaging studies were extracted from the electronic medical record and stored in a REDCap database. De-identified data is available upon request and in compliance with IRB protocols and institutional data sharing agreements.

## ABBREVIATIONS LIST

LR: Likelihood ratio

## Notes

Summary conflict of interest statements: The above authors have no conflicts of interest to report.

Funding information: Jeffrey Sturek is supported in part by the National Center For Advancing Translational Sciences of the National Institutes of Health under Award Numbers UL1TR003015 and KL2TR003016.

### Competing Interest Statement

The authors have declared no competing interest.

### Funding Statement

Jeffrey Sturek was supported in part by the National Center For Advancing Translational Sciences of the National Institutes of Health under Award Numbers UL1TR003015 and KL2TR003016.

### Author Declarations

Study was approved by UVA Health-Sciences Research IRB with approval number 22067. This study is retrospective with a waiver of patient consent.

